# Recent advances in efficacy of corticosteroids as adjunct therapy for the treatment of community-acquired pneumonia in children: a systematic review and meta-analysis

**DOI:** 10.1101/2022.04.25.22274279

**Authors:** Lydia Mukanhaire, Junyan Wang, Xiaoyu Zong, Lingjian Zhang, Xiaohui Zhou, Jian Gong

## Abstract

It has been recently shown that the adjunct use of corticosteroids in the treatment of community-acquired pneumonia shorten the time taken to reach clinical stability (time to clinical stability) in patients with community-acquired pneumonia (CAP). Considering the hyperglycemic effects of corticosteroids, there are concerns about the efficacy and safety of this therapy for children with CAP. Our objective is to evaluate the influence of recent advances in adjunct corticosteroid use and/or aerosolized antibiotic administration on admission to hospital with our main outcome being duration of fever and hospital stay, and additional outcomes as the time to clinical stability therapeutic efficacy, C-reactive protein and defervescence at 24, 48, and 72 hours after starting treatment in a well-defined cohort of children with community-acquired pneumonia. Therapeutic efficacy is defined as the rate of achieving clinical recovery with no fever, improvement or disappearance of cough, and improved or normal laboratory values. Five academic literature databases will be searched using Boolean keyword searches. Articles eligible for inclusion are those that present original research with the study topic as CAP, the study was designed as a randomized controlled trial (RCT) or clinical trial (CT) or an observational study with controls. The review will result in a narrative synthesis that summarizes the effectiveness of corticosteroid use in children.

## Introduction

### Rationale

Pneumonia is an infection that inflames the air sacs in one or both lungs. The air sacs may fill with fluid or pus, causing cough with phlegm or pus, fever, chills and difficulty breathing. A variety of organisms, including bacteria, viruses and fungi can cause pneumonia[1]. Pneumonia can range in seriousness from mild to life threatening[2]. It is most serious for infants and young children, people older than 65,and people with underlying health problems or weakened immune system[3].Pneumonia can be broadly categorized as community-acquired pneumonia, hospital acquired (nosocomial) pneumonia and ventilator associated[4]. Specific pathogens such as mycoplasma pneumoniae, streptococcus pneumoniae, respiratory syncytial virus and haemophilus influenza type b have been found to increase the risk of hospitalization[5].Risk factors for pneumonia in children include infancy, premature birth, incomplete immunization, maternal smoking or household tobacco smoke exposure, indoor air pollution, low birthweight, malnutrition, lack of exclusive breastfeeding and overcrowding [6, 7]. Pneumonia can be spread in multiple ways, the viruses and bacteria that are found in a child’s nose or throat, can infect the lungs if they are inhaled[8]. They may also spread via air-borne droplets from a cough or sneeze. Also, pneumonia may spread through blood during and shortly after birth[9].The presenting features of viral and bacterial pneumonia are similar[10]. However, the symptoms of viral pneumonia may be more numerous than bacterial pneumonia. In children under 5 years of age, who have cough and/or difficult breathing, with or without fever, pneumonia is diagnosed by the presence of either fast breathing or lower chest wall in-drawing where their chest moves in or retracts during inhalation[11]. Wheezing is most common in viral infections. Severely ill infants may be unable to feed or drink and may also experience unconsciousness, hypothermia and convulsions[12]. Pneumonia is treated with antibiotics, amoxicillin being the first line of treatment[13]. Hospitalization is required for severe cases of pneumonia. The most effective way to prevent pneumonia is immunization against Haemophilus Influenza type b (Hib), pneumococcus, measles and whooping cough(pertussis)[14]. Adequate nutrition improves the child’s natural defenses, starting with exclusive breastfeeding for the first six months of life[15]. Encouraging good hygiene and providing affordable clean indoor stoves (in crowded homes) helps to reduce pneumonia infection in children[16]. In children infected with HIV are given cotrimoxazole daily to decrease the risk of contracting pneumonia[17].

### Community-acquired pneumonia

Community-acquired pneumonia (CAP) is the most common type of pneumonia. It occurs outside of hospitals and other healthcare facilities and may be caused by respiratory syncytial virus, streptococcus pneumoniae being the most common bacterial cause, bacteria like organisms like mycoplasma pneumoniae and fungi which comes in soil and bird droppings[18] [19].

### Diagnosis of CAP

#### Clinical Diagnosis

Tachypnoea, hypoxia, cough and increased work of breathing has been found to be signs of pneumonia[20]. Common clinical symptoms of CAP include cough, fever, chills, fatigue, dyspnea, rigors, and pleuritic chest pain. Depending on the pathogen, a patient’s cough may be persistent and dry, or it may produce sputum[21].The World Health Organization criteria in the diagnosis of childhood pneumonia is categorized as follows:

- In children< 2 months :> 60 breaths/minute
- In children 2-12 months:> 50 breaths/minute
- In children > 12 months :>40 breaths /minute

#### Etiological diagnosis

Virus infection, mostly by respiratory syncytial virus is more common in younger children[22]. In older children, the most identified pathogen is streptococcus pneumoniae, followed by mycoplasma and chlamydia[23]. Diagnostic modalities available for etiological diagnosis include molecular diagnostics, microscopy, culture and antigen detection[24]. Both bacterial and viral pneumonia exhibit a wide distribution of acute phase reactants (blood count, C reactive protein, erythrocyte sedimentation rate)[22].Upper respiratory tract secretions are useful in virological diagnosis. Pulmonary TB should be considered in a child presenting with severe pneumonia or pneumonia with a known TB contact[25].

#### Radiological diagnosis

The radiological signs of pneumonia overlap with those of collapse. Chest radiography does not distinguish between viral and bacterial infection and is unable to detect early changes in pneumonia.[7]However, chest radiography improves the diagnosis of pediatric community acquired pneumonia to a certain degree and may prevent over-treatment with antibiotics[26].

#### Assessment of severity

The general appearance of a child should be assessed in order to evaluate the severity of the illness. Any general danger sign requires a child to be referred to a hospital[27].All children less than 2 months of age with signs of pneumonia require hospital admission. Inability to drink/feed, vomiting everything, convulsions, lower chest in-drawing, central cyanosis, lethargy, nasal flaring, grunting, head nodding, and oxygen saturation <90% are predictors of death and can be used as indicators for hospitalization[28].Pulse oximetry should also be performed in all children, using a pediatric wrap-around probe. A saturation of<92% or 90% at higher altitudes (>1 800m) indicates the need for hospital admission and supplemental oxygen[29]. Transfer to pediatric intensive care unit should be considered when: the patient is shocked, failure to maintain rising respiratory and pulse rates with clinical evidence of severe respiratory distress, recurrent apnea or slow irregular breathing[30]

### Corticosteroid

Corticosteroids are synthetic analogs of the natural steroid hormones produced by the adrenal cortex and include glucocorticoids and mineral corticoids[31]. Corticosteroids work primarily by modulating transcriptional, post-transcriptional, and post-translational mechanisms within cellular nuclei to decrease the production of inflammatory mediators[32]. Glucocorticoids are involved in metabolism and have immune-suppressive, anti-inflammatory and vasoconstrictive effects while mineral corticoids regulate electrolytes and water balance by affecting ion transplant in the epithelial cell of renal tubules[33]. Corticosteroids are readily absorbed in the gastrointestinal tract and are highly protein-bound. They undergo hepatic metabolism and renal excretion[31]. Corticosteroids at high concentrations will also inhibit the production of B cells and T cells[34].

### How intervention might work

With the increasing reported cases of pneumonia in recent years, there has been an introduction of other supportive therapies for the treatment of MP infection[35]. Adjunct treatment therapies for CAP infections include the use of corticosteroids and aerosolized antibiotics[36].This might be due to the rapidly increased cases among children, particularly sub-Saharan Africa and in East Asian countries such as Korea, Japan, and China. As Corticosteroids have been shown to be of use as adjunct therapy in the treatment of community-acquired pneumonia (CAP) it is important to note their effects and physiological roles. The clinical implications of these therapies have not been fully elucidated[37], thus it is imperative to note any advancements in these therapies. Moreover, there is need for supportive treatment options in CAP cases with clinical deterioration. However, the use of corticosteroids in children is limited because of their toxicity. Methylprednisolone may cause adverse effects such as growth retardation, weight gain, Cushingoid features and osteoporosis in children[38, 39]. Therefore, they are contraindicated for premature infants. Aerosolized antibiotics have been found to provide high concentration of drugs at the target site of action in patients with pulmonary infections[17].

## Methods

### Search strategy

Our search strategy is developed based on systematic review best practices. To identify relevant studies, we will perform an extensive search across 5 electronic full-text databases; Medline/PubMed, Embase, the Cochrane Library, Scopus and Web of Science with no language restrictions. These databases will be searched using keywords for community acquired pneumonia, corticosteroid and children as shown in a table (Table 2). Database specific Boolean search strategies were developed and follow the general format: corticosteroid terms AND (community acquired pneumonia terms or children’s terms) we will search articles published from January 1967 to March, 2022 using a protocol designed for this study. The search terms used for each database are listed below:

**Table 1.** Databases included in the systematic review.

| Databases | URL |
| --- | --- |
| Web of science | www.webofknowledge.com |
| Medline/PubMed | pubmed.ncbi.nlm.nih.gov/ |
| Embase | www.embase.com |
| Scopus | www.elsevier.com |
| The Cochrane Library | www.cochranelibrary.com/ |
| World Health Organization | www.who.int/health-topics/pneumonia |

**Table 2.** Search keywords.

| Corticosteroid terms | Children terms |
| --- | --- |
| “Corticosteroids” | “Pediatrics” |
| “Prednisolone” | “Infant” |
| “Cortisone” | “Children” |
| “Hydrocortisone” | “Pre-school child” |
| “Budesonide” | “Child” |
| “Dexamethasone” | “Newborn” |
| “Prednisone” | “Toddler” |
| “Glucocorticoids” | “Kid” |
| “Mineral corticoids” | “Neonate” |
| **Pneumonia terms** |  |
| “Community-acquired pneumonia” |  |
| “Mycoplasma pneumonia” |  |
| “Pediatric-pneumonia” |  |
| “Childhood-pneumonia” |  |

### Study selection, quality assessment, and data extraction

Studies are to be screened by two independent reviewers based on the title and abstract, followed by full-text screening. The literature selection process will be conducted in accordance with the Preferred Reporting Items for Systematic Reviews and Meta-Analysis Protocols 2015 statement[40]. The quality of the selected studies will be assessed using the Cochrane Risk of Bias Tools for RCTs and the revised Risk of Bias Tool for Non-Randomized Studies for observational studies. The data extraction form will include the following information: first author, year of publication, population in each group, antibiotic treatment (macrolides/comparator), patient characteristics (age and sex), and outcomes (durations of fever and hospitalization, therapeutic efficacy, and defervescence rates at 24, 48, and 72 h after starting treatment). Therapeutic efficacy is defined as the rate of achieving clinical recovery with no fever, improvement or disappearance of cough, and improved or normal laboratory values. Study selection, quality assessment, and data extraction to be conducted. Any disagreements to be resolved through discussion. If the results of the selected studies are unclear or missing, we will contact the corresponding study investigators to obtain or confirm data

### Eligibility criteria

Articles that meet the following inclusion criteria will be included: (1) the study topic was CAP, defined as a disease showing no clinical or radiological improvement 48– 72 h after macrolide administration; (2) the subjects are children aged ≤ 12 years (3)the study was designed as a randomized controlled trial (RCT), Clinical Trial (CT) or an observational study with controls; (4) the intervention agent should be a corticosteroid known to be active against community-acquired pneumonia, such as methylprednisolone (5) the control was a placebo; and (6) at least one of the predetermined outcomes was reported. Animal and preclinical studies, as well as articles other than original research articles (e.g., reviews, editorials, letters, conference abstracts, and comments) are to be excluded. Studies with duplicate subjects (i.e., different studies using the same outcome indicators in the same number of patients) are also going to be excluded. Our search strategy will implement no language restrictions, and non-English articles should be translated and included for evaluation

### Data synthesis

Statistical synthesis of data(meta-analyses) will be conducted. A systematic narrative synthesis will be provided with the information presented in text and tables to summarize and explain the characteristics and findings of the included studies.

The following is a tentative outline of how we will synthesize findings. First, community-acquired pneumonia in children’s conceptualizations will be summarized. This will include definitions provided by community-acquired pneumonia researchers. Second, the antecedents of community-acquired pneumonia in children will be summarized. This will likely to include the grouping of corticosteroid therapies in the treatment of community-acquired pneumonia in children from literature. Finally, the evidence on recent advances in efficacy of corticosteroids and other supportive therapies in the treatment of community-acquired pneumonia will be incorporated into the theoretical framework.

### Statistical analysis

We will pool the findings from the included studies such as calculated mean, standard deviation, and sample size. For outcomes presented as continuous variables, such as fever duration, hospital stay length, and therapeutic efficacy, we will calculate mean differences with 95% confidence intervals (CIs). For dichotomous outcomes such as the achievement of defervescence after 24, 48, and 72 h of treatment, we will calculate odds ratios (ORs) with 95%CIs. The average effect summary will be calculated using a random-effects model (Mantel–Haenszel method) using Review Manager 5.3 (The Cochrane Collaboration, London, UK) including the *I*2 statistic. *I*2 of 25%, 50%, and75% indicated low, moderate, and high heterogeneity, respectively. To assess the risk of publication bias, we will use funnel plots for visual inspection; Egger test flow chart of the selection process to be included in meta-analysis. The strength of the body of evidence will be assessed using GRADE approach[41].

### Flow chart of the selection process of studies to be included in the meta-analysis

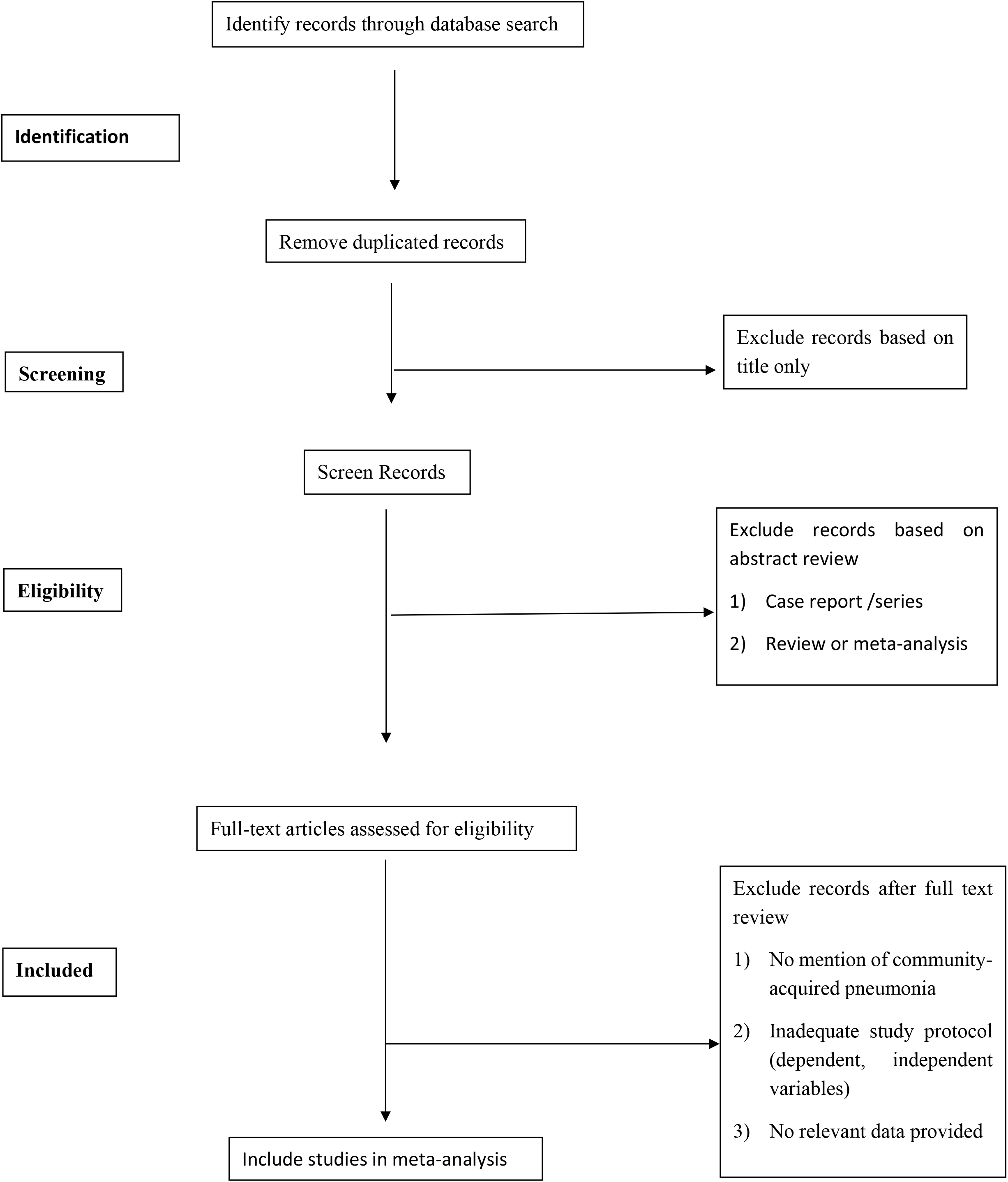

### Risk of publication bias

The authors have no conflicts of interest. Ideally, additional reviewers would participate in study screening and data extraction. However, this is not feasible since this publication is not funded.

### Ethics and dissemination

This systematic review protocol has been written following the PRISMA-P guidelines and elaboration[40, 42].

## Discussion

Corticosteroids may be imperative for the treatment of community-acquired pneumonia. Therapeutic doses vary greatly, as do adverse effects. Patients require education on what to expect with corticosteroid use, whether it be short course or long-term use. Other pharmacological therapies may be necessary to counteract corticosteroid-related adverse effects, such as gastric acid suppression, calcium and vitamin D supplementation, and opportunistic infection prophylaxis. Providers must weigh the risks versus benefits of corticosteroid use and utilize the lowest effective dose for the least duration possible to avoid or minimize serious corticosteroid-induced toxicities. This review searches for the efficacy of corticosteroids as adjunct therapy in the treatment and management of community-acquired pneumonia in children. There is an ongoing unclear elucidation on the effectiveness of corticosteroids use specifically in children under the ages of 5. By carefully examining the published literature to date, we will contribute a novel synthesis of literature compared to “corticosteroid use in community-acquired pneumonia” literature that has been published before.

## Data Availability

All relevant data from this study will be made available upon study completion.

## Supporting information

1. Table .PRISMA-P checklist (Docx)
2. Search strategy draft(doc)

## Author Contributions

**Conceptualization:** Lydia Mukanhaire, Jian Gong

**Methodology:** Lydia Mukanhaire, Jian Gong

**Supervision:** Xiaohui Zhou, Jian Gong

**Writing-original draft:** Lydia Mukanhaire

**Writing-review & editing:** Junyan Wang, Xiaoyu Zong, Lingjian Zhang

